# More tests, higher scores: Testing frequency and SDMT differences between natalizumab and rituximab

**DOI:** 10.64898/2026.10.21.26366102

**Authors:** Timothy P. Copeland, Sanabel Abdel Hay, Fredrik Piehl, Katharina Fink, Kyla A. McKay

**Affiliations:** Department of Clinical Neuroscience, Karolinska Institutet, Stockholm, Sweden; Center for Molecular Medicine, Karolinska University Hospital, Stockholm, Sweden

**Keywords:** multiple sclerosis, cognition, Symbol Digit Modalities Test, natalizumab, rituximab, practice effects, observational studies

## Abstract

**Background:** The Symbol Digit Modalities Test (SDMT) measures information processing speed, and repeated testing can increase scores through practice effects. However, how differences in testing frequency bear on comparisons between disease-modifying therapies (DMTs) in real-world cohorts has not been fully explored.

**Objective:** To compare SDMT trajectories between natalizumab and rituximab users and quantify the association of the testing process with those trajectories.

**Methods:** Using the Swedish MS Registry, we studied adults with relapsing-remitting MS initiating natalizumab or rituximab (2010–2024) with ≥2 on-treatment SDMT assessments. Mixed-effects models with restricted cubic spline time were sequentially adjusted; log cumulative test number was the practice-effect adjustment.

**Results:** Among 2,458 individuals, natalizumab initiators were tested twice as often as rituximab initiators (1.6 vs. 0.8 tests/year). The predicted natalizumab-minus-rituximab SDMT difference at 3 years was 7.0 points (95% CI 5.7–8.3) unadjusted and 1.6 (0.7–2.6) after full adjustment including cumulative test number; at 1 year it fell from 4.4 (3.3–5.4) to −0.3 (−1.2 to 0.6).

**Conclusion:** Adjustment for cumulative test number substantially attenuated the SDMT difference between natalizumab and rituximab, leaving a small residual difference of uncertain clinical importance and underscoring the need to account for testing frequency in real-world comparisons.

## Introduction

Cognitive dysfunction is among the most disabling features of multiple sclerosis (MS)^1, 2^. Among people with MS (PwMS), 30–45% of those with relapsing-remitting disease show measurable impairment^1^, often emerging within the first years of disease and progressing largely independently of physical disability^2^. Information processing speed, measured by the Symbol Digit Modalities Test (SDMT), is a highly sensitive and reliable cognitive marker in MS and a core component of the Brief International Cognitive Assessment for MS^3, 4^.

Repeated administration of the same cognitive test inflates performance through familiarity, known as a practice effect^5^. For the SDMT, scores typically increase after the first administration^6^, before plateauing^7^. Altering the symbol-digit key reverses much of that gain, which implies that test takers are learning the testing key^8^. The practice-effect contribution therefore depends on cumulative tests. In routine care, cognitive testing is often tied to clinic visits, so for infusion disease-modifying therapies (DMTs) given on-site the dosing interval sets how often patients are tested.

Natalizumab and rituximab are high-efficacy infusion DMTs for relapsing-remitting MS^9–11^. Natalizumab is administered every 4 to 6 weeks on average^12^, while rituximab is typically given every 6 months or at longer 12-month intervals^11, 13^.

Previous observational studies have suggested better cognitive outcomes with natalizumab compared with rituximab and other DMTs. A cross-sectional multicentre study found SDMT improvement from initiation on natalizumab but not rituximab (+8.9 vs. −1.0 points, p = 0.002)^14^, and a nationwide registry analysis reported higher odds of SDMT improvement on natalizumab than on other DMTs (odds ratio 2.3, 95% CI 1.5–3.5)^15^. A group-based trajectory analysis found processing speed broadly stable after DMT start without estimating drug-specific trajectories^16^. Most recently, researchers reported that high-efficacy DMTs were associated with an initial increase followed by a subsequent decline in SDMT scores, with the early increase attenuated after adjustment for the number of repeated SDMT measurements^17^. None of these designs separated accumulated testing as it accrues over follow-up (i.e., practice effects) from treatment-related cognitive change. Here, we compare SDMT trajectories between natalizumab and rituximab in a nationwide MS registry cohort and quantify the association of the testing process with those trajectories.

## Methods

### Study design and setting

We conducted a retrospective longitudinal cohort study using prospectively collected data from the Swedish MS Registry (SMSreg), a nationwide registry covering approximately 86% of PwMS in Sweden^18^. The registry contains demographics, MS phenotype, DMT episodes, Expanded Disability Status Scale (EDSS) scores, clinician-recorded relapses, and SDMT assessments. A previous validation study involving chart review of more than 3,000 patients demonstrated the overall reliability of registry data, although completeness varies across clinical variables and patients^19^. Systematic SDMT collection began through the Immunomodulation and Multiple Sclerosis Epidemiology (IMSE) natalizumab post-marketing surveillance programme in 2006 and was subsequently extended to all newly approved DMTs^15^.

### Study population

Individuals with a first recorded natalizumab or rituximab episode in the SMSreg (with no prior exposure to the comparator) between 1 January 2010 and 31 December 2024 were included. Eligibility also required a recorded relapsing-remitting subtype, age ≥18 at index with a known date of birth, a valid baseline SDMT within 0–90 days before DMT start, no SDMT testing before the baseline window, and ≥2 SDMT assessments during the on-treatment episode. The cohort flow appears in Supplementary Figure S1; Supplementary Table S1 compares the 2,458 included with the 6,602 excluded on SDMT criteria among 9,060 otherwise eligible.

### Exposure, outcome, and follow-up

The exposure was the index DMT, natalizumab or rituximab. Follow-up began at treatment initiation and continued until treatment discontinuation, death, loss to follow-up, or the study end (31 December 2024), whichever occurred first. Only SDMT assessments recorded during this on-treatment follow-up period were included in the analysis. The primary outcome was the SDMT score, the number of correct symbol-digit matches made in 90 seconds using a printed reference key, modelled as a continuous repeated measure^3^. Equivalent alternate SDMT forms exist^20^, and Swedish guidance recommends alternating versions to mitigate practice effects from repeated testing^21^. The IMSE programme distributed alternate SDMT forms annually, although clinics report using only the original version in routine care. The oral SDMT was used nationwide, except in Stockholm, which switched from written to oral administration after 2017 (K. Fink, personal communication, July 2026). Rituximab was not included in IMSE until the prospective phase of COMBAT-MS began in 2017, when participating clinics were instructed to test once yearly and to use oral administration only (F. Piehl, personal communication, October 2026). Only scores between 5 and 110 points were included.

### Practice effects and test-number adjustment

The variable of interest was cumulative SDMT test number, counted from and including baseline. Its natural log was entered as a time-varying covariate to absorb the upward drift from repeated administration while letting the marginal increment diminish, consistent with gains that slow and plateau with repetition^7, 8^. Baseline SDMT was not modelled as an outcome but entered as a covariate, so adjusted trajectories compare on-treatment SDMT at a common baseline. This adjustment would isolate practice effects only if testing times and test number varied independently of treatment. Because treatment-linked monitoring largely determines testing frequency, this does not hold true. We therefore treat the practice-adjusted estimate as an upper bound on a possible treatment effect rather than as a corrected estimate, assuming the log test-number term absorbs practice gain and no treatment-related gain; without that assumption, only the unadjusted contrast bounds the treatment effect.

### Covariates

Baseline covariates measured at index were age, sex, disease duration, EDSS^22^ (nearest value within 180 days before index), treatment-naïve status, calendar year, health care region recorded in SMSreg, and SDMT. The time-varying covariate was cumulative SDMT test number. Missing baseline EDSS and disease duration were filled by single stochastic imputation conditional on age, sex, and region (Supplementary Methods).

### Statistical analysis

Baseline characteristics were summarised by treatment using means with SD, medians with Q1 and Q3, and counts with percentages. Trajectory shape was evaluated by comparing linear time with restricted cubic spline (RCS) time using 3, 4, and 5 knots at Harrell’s recommended percentiles^23^, fitted unadjusted by maximum likelihood and compared by Akaike Information Criterion (AIC); the specification with the lowest AIC became the primary time structure (Supplementary Table S2, Supplementary Figure S2, Supplementary Methods). We fitted mixed-effects models with random intercepts and random linear slopes for time, an unstructured covariance for the random effects, and robust standard errors. Sequential adjustment proceeded through three nested models: (1) unadjusted; (2) adjusted for baseline covariates; and (3, primary) plus log-transformed cumulative test number. Calendar year entered as a three-knot restricted cubic spline interacted with time, so that calendar period could alter the slope as well as the level of the trajectory, and region as fixed effects; regions with fewer than 20 individuals in either arm, and missing region, were pooled into one category; no region met this criterion and no region was missing, so all six health care regions entered separately. Individual spline coefficients have no standalone interpretation, so we summarised each model by the predicted natalizumab-minus-rituximab difference at 1, 2, and 3 years from the fitted curves and by the initial slopes, which are linear until the first knot of the spline and cover the earliest months of treatment (i.e., up to the 5th percentile of follow-up time). Full coefficients are given in Supplementary Table S3. We also evaluated the primary model with test number set to the count reached by each time point on each arm’s observed schedule (a lowess of test number on time, fitted by arm), including natalizumab on the rituximab schedule and displayed the results graphically.

We asked how well treatment could be predicted from three feature groups: how a person was tested (on-treatment SDMT count, testing rate, intervals between tests, and timing of the first and last test), their baseline clinical characteristics (age, sex, EDSS, disease duration, treatment-naïve status, and baseline SDMT), and calendar year of initiation. If ‘how a person is tested’ alone can identify the drug, then we can conclude that testing and treatment cannot be separated in the trajectory models. We fitted gradient-boosted classifiers^24^ to each group and to all three combined, summarising discrimination by the five-fold cross-validated area under the receiver operating characteristic curve (AUC; Supplementary Table S4) and ranking feature contributions using SHapley Additive exPlanations (SHAP) values (Supplementary Table S5). Feature definitions, settings, and diagnostics are in the Supplementary Methods.

Analyses were conducted in Stata 19 and Python (scikit-learn).

### Ethics

The Regional Ethical Review Board in Stockholm approved the study (2017/1378-31). Individual informed consent was waived because the study used pseudonymised registry data.

## Results

### Cohort and baseline characteristics

Of 13,649 people with a first natalizumab or rituximab episode, 2,458 met all eligibility criteria (Supplementary Figure S1).

PwMS initiating natalizumab were younger than those initiating rituximab, more often had prior DMT exposure, and had greater upper-quartile baseline EDSS (Table 1). Natalizumab was also initiated earlier in calendar time (median index year 2013 vs. 2018). Sex distribution, disease duration, and baseline SDMT score were comparable. Before imputation, baseline EDSS was missing for 956 individuals (38.9%) and disease duration for 52 (2.1%). PwMS on natalizumab had a median of 6 on-treatment SDMTs versus 5 on rituximab and twice the testing rate (median 1.6 vs. 0.8 SDMTs/year), despite shorter time to the last on-treatment SDMT (median 41.5 vs. 55.2 months). Figure 1 shows the schedule asymmetry visit by visit, the first three on-treatment tests clustering within about 1.5 years for natalizumab and extending to roughly 2.5 to 3 years for rituximab.

**Table 1.** Baseline characteristics of the study cohort, by treatment group, including the number with baseline EDSS and disease duration missing before imputation.

| **No. (Column %), Mean±SD, or Median (Q1, Q3)** | **Total**<br>**N=2,458** | **Rituximab**<br>**N=1,384** | **Natalizumab**<br>**N=1,074** |
| --- | --- | --- | --- |
| Age at treatment initiation (years) | 36.4±10.4 | 38.3±10.7 | 34.0±9.4 |
| Sex |  |  |  |
| Male | 753 (31) | 436 (32) | 317 (30) |
| Female | 1,705 (69) | 948 (68) | 757 (70) |
| Disease duration at treatment (years) | 2.4 (0.4, 7.8) | 2.3 (0.4, 8.6) | 2.4 (0.5, 7.0) |
| Disease duration |  |  |  |
| <2 years | 1,146 (47) | 655 (47) | 491 (46) |
| 2-5 years | 449 (18) | 235 (17) | 214 (20) |
| >5 years | 863 (35) | 494 (36) | 369 (34) |
| Baseline EDSS | 2.0 (1.0, 2.6) | 2.0 (1.0, 2.5) | 2.0 (1.0, 2.8) |
| Baseline EDSS missing before imputation | 956 (39) | 562 (41) | 394 (37) |
| Disease duration missing before imputation | 52 (2) | 39 (3) | 13 (1) |
| Baseline SDMT score | 51.8±11.4 | 51.8±10.9 | 51.8±11.9 |
| Cognitively impaired at baseline (SDMT <45) | 594 (24) | 333 (24) | 261 (24) |
| Treatment naive (no prior DMT) | 1,391 (57) | 909 (66) | 482 (45) |
| Duration of prior DMT exposure (months) | 0.0 (0.0, 25.8) | 0.0 (0.0, 20.0) | 4.2 (0.0, 30.8) |
| Number of on-treatment SDMT tests | 5.0 (3.0, 8.0) | 5.0 (3.0, 6.0) | 6.0 (3.0, 12.0) |
| Years of observation | 5.5 (3.1, 7.8) | 6.0 (4.2, 7.7) | 4.1 (2.1, 7.8) |
| SDMT measurements per year | 1.0 (0.7, 1.6) | 0.8 (0.6, 1.1) | 1.6 (1.1, 2.1) |
| Calendar year of initiation | 2016 (2013, 2019) | 2018 (2016, 2019) | 2013 (2011, 2016) |

*Footnote:* EDSS, Expanded Disability Status Scale; SDMT, Symbol Digit Modalities Test; DMT, disease-modifying therapy; Q1 and Q3, first and third quartiles. Baseline EDSS and disease duration include values filled by single stochastic imputation (Supplementary Methods), so EDSS quartiles need not fall on EDSS steps.

**Figure 1.**
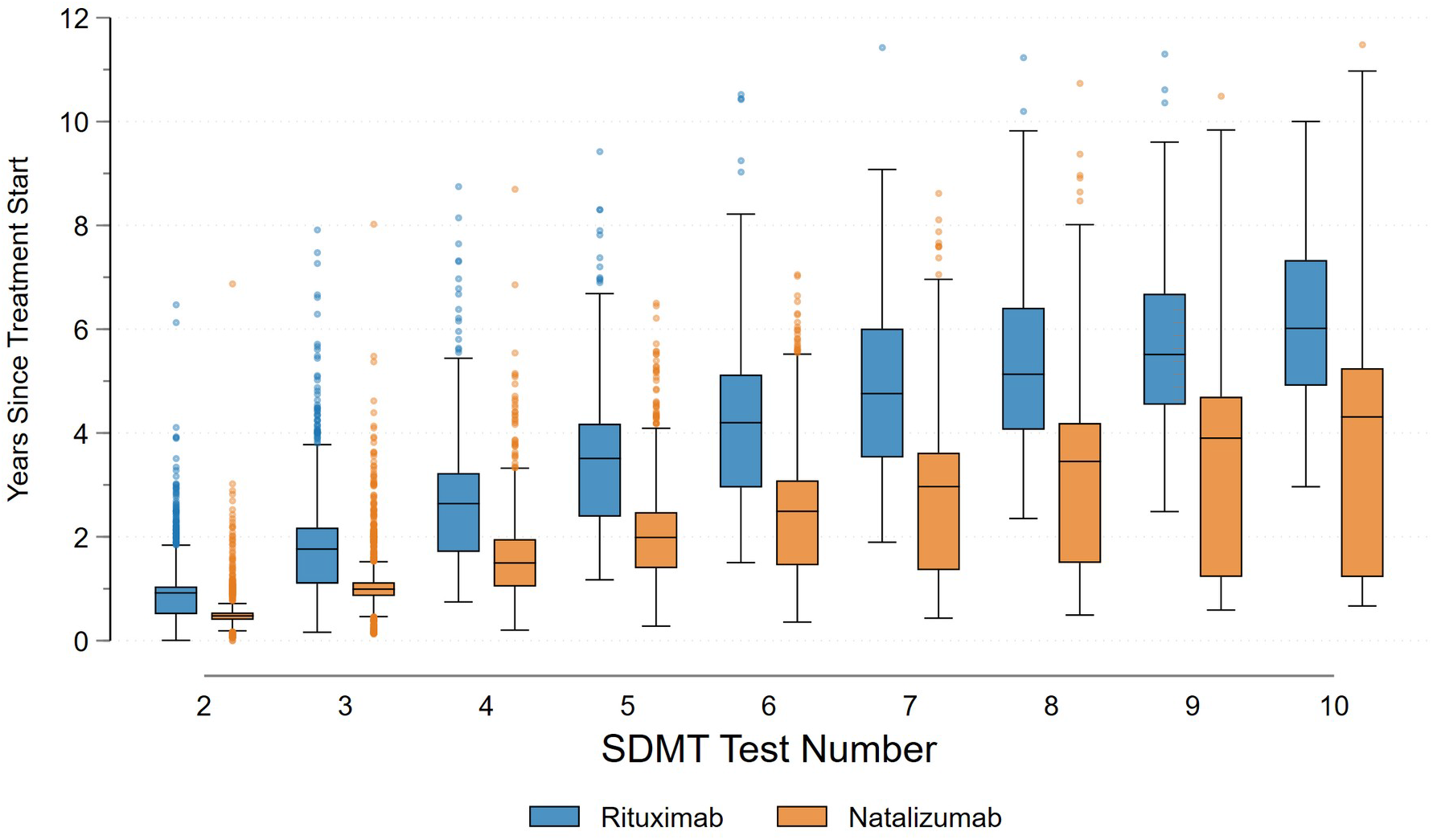
Timing of successive SDMT assessments by treatment group. Box plot of years since treatment initiation at successive on-treatment SDMTs (tests 2 to 10), by treatment arm; markers show outlier values rather than every visit. Test numbers count from the baseline (pre-treatment) assessment, so test 2 is the first on-treatment SDMT.

### Unadjusted SDMT outcomes

The cohort contributed 18,300 on-treatment SDMT assessments (mean score 60.1, SD 15.9). Descriptive change scores favoured natalizumab on every metric (Table 2). The mean change from baseline to last SDMT was 7.8 (SD 12.2) points among natalizumab initiators versus 3.1 (SD 8.3) among rituximab initiators, and last-visit SDMT was 59.6 vs. 54.9 from indistinguishable baselines.

**Table 2.** SDMT outcomes during follow-up, by treatment group.

| **No. (Column %), Mean±SD, or Median (Q1, Q3)** | **Total**<br>**N=2,458** | **Rituximab**<br>**N=1,384** | **Natalizumab**<br>**N=1,074** |
| --- | --- | --- | --- |
| Number of on-treatment SDMT tests | 5.0 (3.0, 8.0) | 5.0 (3.0, 6.0) | 6.0 (3.0, 12.0) |
| Time to last on-treatment SDMT (months) | 50.5 (28.2, 76.8) | 55.2 (35.1, 74.8) | 41.5 (22.6, 82.8) |
| Baseline SDMT score | 51.8±11.4 | 51.8±10.9 | 51.8±11.9 |
| Last SDMT score | 56.9±14.1 | 54.9±12.5 | 59.6±15.6 |
| Change: baseline to last SDMT | 5.2±10.5 | 3.1±8.3 | 7.8±12.2 |
| Mean SDMT change from baseline | 4.4±8.4 | 2.4±6.6 | 6.9±9.6 |
| Maximum SDMT change | 10.0±10.4 | 7.3±7.9 | 13.4±12.0 |
| Any recent relapse (within 120 days of an on-treatment SDMT) | 141 (6) | 27 (2) | 114 (11) |

*Footnote:* SDMT, Symbol Digit Modalities Test; Q1 and Q3, first and third quartiles.

### Longitudinal trajectories depend on the practice-effect specification

Among the candidate time specifications, AIC favoured the five-knot spline (Supplementary Table S2). On the fitted five-knot curves, the predicted natalizumab-minus-rituximab difference widened from 4.4 points at 1 year to 6.6 at 2 years and 7.0 at 3 years unadjusted, and to 1.6, 3.6, and 3.7 after baseline covariate adjustment. Adding log cumulative test number (Table 3) cut each of these to −0.3 (95% CI −1.2 to 0.6), 1.0 (0.1–1.9), and 1.6 (0.7– 2.6). A similar pattern emerged in the linear treatment-by-time covariate (i.e., the slope until the first knot at the 5th percentile of time); over the first months of treatment, before the bend in the spline, natalizumab-treated PwMS improved 5.28 points/year (3.85–6.71) faster than rituximab-treated PwMS unadjusted. However, when the practice term was added, this difference fell to 1.38 (0.18–2.57; Supplementary Table S3). The practice term itself (log-transformed cumulative test number) was large and precise: 6.60 points per log unit of cumulative test number (5.35–7.86), or 4.6 points (3.7–5.4) per doubling.

**Table 3.** Predicted natalizumab-minus-rituximab difference in SDMT at 1, 2, and 3 years under sequential adjustment.

| **Years since initiation** | **Unadjusted** | **Adjusted** | **+ Practice (primary)** |
| --- | --- | --- | --- |
| Year 1 | 4.35 (3.31, 5.39) | 1.56 (0.76, 2.36) | -0.31 (-1.17, 0.55) |
| Year 2 | 6.63 (5.44, 7.82) | 3.60 (2.67, 4.53) | 1.00 (0.10, 1.89) |
| Year 3 | 7.01 (5.74, 8.27) | 3.70 (2.67, 4.73) | 1.62 (0.65, 2.60) |

*Footnote:* Values are predicted differences in SDMT points with 95% confidence intervals from robust standard errors, from restricted cubic spline (five-knot) mixed-effects models of longitudinal SDMT trajectories.
Adjusted: age, sex, baseline EDSS, disease duration, treatment-naïve status, baseline SDMT, calendar year of initiation as a three-knot restricted cubic spline with each spline term interacted with time since initiation, and health care region as fixed effects (regions with fewer than 20 individuals in either arm, and missing region, pooled). + Practice (primary): adjusted plus log cumulative test number. Predicted differences are evaluated on the fitted five-knot curve at 1, 2, and 3 years with covariates at their means, log test number at its sample mean in the + Practice model, and region as observed.

Natalizumab, on its own testing schedule, climbs steeply over the first two years and sits roughly 10 points above rituximab at 3 years, whereas natalizumab moved onto the rituximab schedule stays within 2 points of rituximab (Figure 2), representing the primary contrast in Table 3.

**Figure 2.**
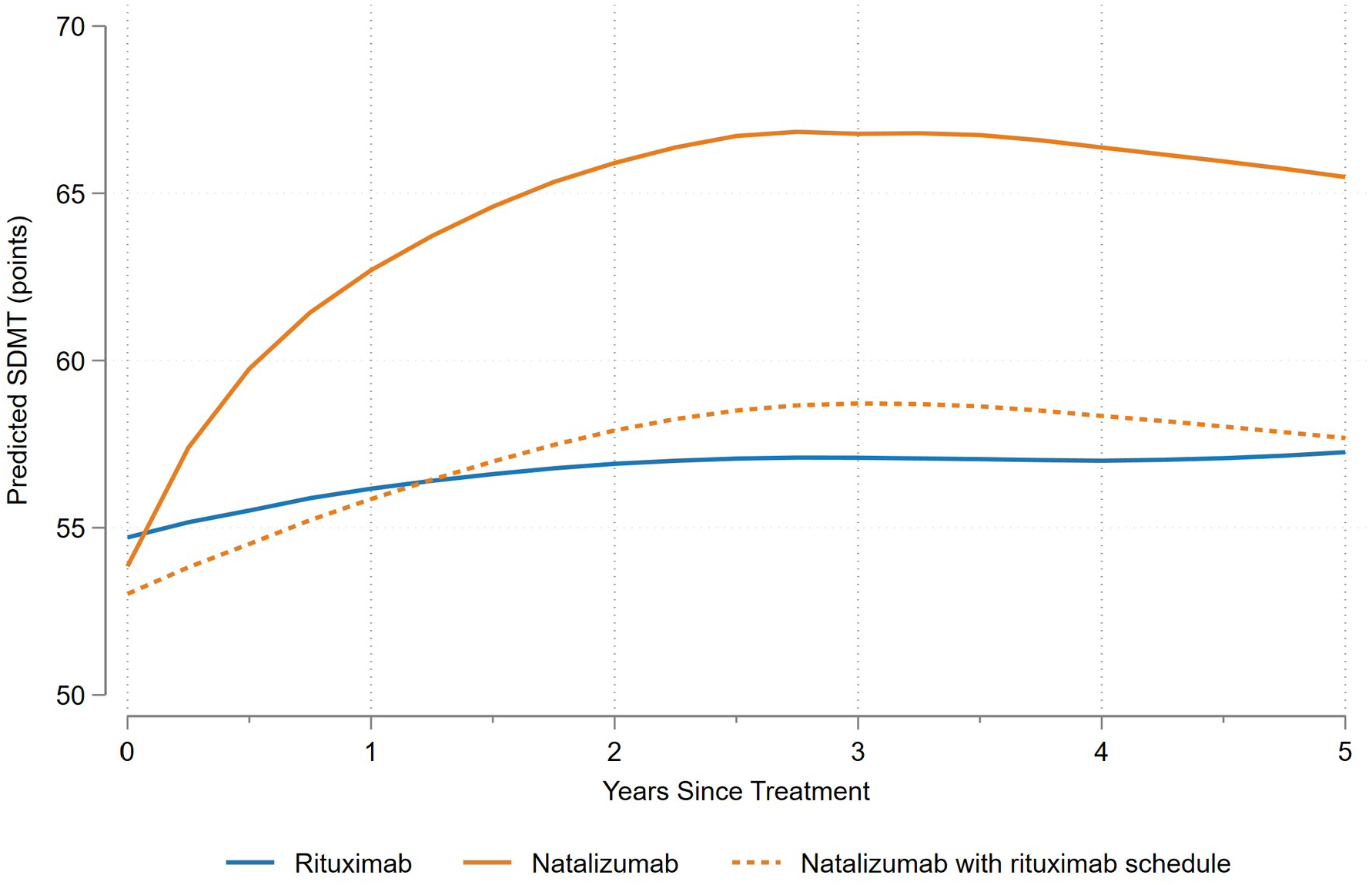
Adjusted SDMT trajectories under each treatment arm’s testing schedule. Predicted SDMT over 0–5 years from the primary five-knot spline model (Table 3, + Practice), with cumulative test number set to the count a patient reaches by each time point: rituximab on the rituximab schedule, natalizumab on the natalizumab schedule, and natalizumab on the rituximab schedule (dashed). The expected test count by arm is a lowess of cumulative test number on time since initiation, fitted separately by arm and floored at the baseline test. Other covariates are held at their means and region as observed.

Adding the practice term also removed the curvature the spline had fit. Without it, the initial time slope for rituximab was positive (1.96 points/year, 1.17–2.75) and the natalizumab-by-spline terms were large and significant; with the practice term, that initial slope turned negative (−2.72, −3.83 to −1.62) and no spline or treatment-by-spline term remained significant (all p > 0.2; Supplementary Table S3). Following inclusion of the practice term, the treatment-by-spline coefficients shrank several-fold, the largest by more than an order of magnitude, while confidence-interval widths stayed comparable, and AIC improved. Because the primary model contains both time and cumulative test number, the two can be examined separately. When every patient is given an equal number of tests, predicted SDMT fell gradually over five years on both natalizumab and rituximab. Time on treatment alone was not associated with higher SDMT in either arm (Supplementary Figure S3A). When time is held fixed instead of cumulative test number, in a separate model that replaces the treatment-by-time interaction with a treatment-by-log-test-number interaction, predicted SDMT rose with each subsequent test for both natalizumab-and rituximab-treated PwMS (i.e., 52.6 to 59.7 points between tests 2 and 10 on rituximab and from 51.7 to 63.7 on natalizumab; Supplementary Figure S3B, Supplementary Table S6). These equal-test-number comparisons rely on the few patients with comparable test counts in both arms and are not estimates of the treatment differences.

### Prediction of treatment assignment

Treatment (rituximab or natalizumab) was highly predictable from how a person was tested. Testing-process features alone reached a cross-validated AUC of 0.88, against 0.68 for baseline clinical characteristics, 0.82 for calendar year, and 0.92 for all features combined (Supplementary Table S4) in predicting treatment. Calendar year and the leading testing-schedule features outranked every baseline clinical variable in the feature ranking (Supplementary Table S5).

## Discussion

In this nationwide cohort, natalizumab-treated PwMS showed greater increases in SDMT scores than rituximab-treated PwMS and were tested approximately twice as often. The predicted between-treatment difference at three years decreased from 7.0 points unadjusted to 3.7 after baseline covariate adjustment and to 1.6 after additional adjustment for cumulative test number. These findings suggest that differences in testing frequency may contribute substantially to apparent differences in cognitive outcomes between DMTs in routine care.

The adjusted three-year difference of 1.6 SDMT points was below the commonly used four-point benchmark for clinically meaningful change^3^, although this benchmark should be applied cautiously to mean differences between treatment groups. Our findings suggest that differential testing frequency may contribute to the natalizumab advantage previously reported in Swedish cohorts^14, 15^.

The most recent Swedish registry study of SDMT differences between natalizumab and anti-CD20 therapies found a steeper early rise and more pronounced decline in the natalizumab arm^17^. That study entered the number of repeated SDMT measurements among its baseline covariates, a person-level adjustment that shifts the level of each trajectory but not its slope, whereas we modelled cumulative test number as a time-varying covariate. Trajectory estimates may also depend on the specification of time: quadratic models constrain curvature, whereas restricted cubic splines allow more flexible shapes. We compared linear and spline specifications using AIC and selected the best-fitting model among those evaluated.

The prediction analysis further highlighted the strong association between treatment and testing frequency. Based solely on SDMT testing patterns, natalizumab- and rituximab-treated individuals could be correctly distinguished in 88% of randomly selected pairs, illustrating the difficulty of separating treatment-related differences from testing practices.

Prior work supports a practice effect of this size and shape. Altering the symbol-digit key reverses much of the accumulated gain^8^, and longitudinal data show practice effects build across administrations^6^ before plateauing^7^. Smartphone self-administered SDMT gives an estimate not tied to treatment-driven testing schedules: in Floodlight Open, e-SDMT practice effects approached a modelled boundary improvement of 40.8% (95% CI 39.9–41.6) over baseline. Half of the practice effect was reached by the 11th repetition and 90% by the 35th, a flattening consistent with the log test-number specification used here^25^. In the ASCEND trial, people with secondary progressive MS improved by a mean of 4.5 SDMT points over 96 weeks regardless of treatment assignment, a gain the investigators attributed to practice^26^.

A biological contribution to the observed SDMT difference cannot be excluded, although greater suppression of inflammatory disease activity by natalizumab is not an obvious explanation. Randomized head-to-head comparisons with rituximab are lacking, but the recent prospective MultipleMS study of newly diagnosed MS found the lowest relapse rate with B-cell-depleting therapies, a reduction in T2 lesion volume with B-cell-depleting therapies and a similar trend with other high-efficacy therapies (a group that included natalizumab), and no adjusted differences in serum neurofilament light chain at 24 months^27^. Although these were grouped treatment comparisons, the findings do not support superior suppression of inflammatory activity as an explanation for the apparent natalizumab cognitive advantage.

### Strengths and limitations

Strengths include a large nationwide cohort with prospectively recorded SDMT assessments, flexible modelling of time, and adjustment for cumulative test exposure as a time-varying covariate. We also assessed how well testing patterns alone predicted treatment, providing an additional measure of the association between DMT use and the testing process.

Several limitations should be considered. The observational design leaves scope for residual confounding despite covariate adjustment, particularly given differences in age and calendar period between treatment groups. Because natalizumab was mostly initiated earlier (median index year 2013 vs. 2018), the calendar-by-time terms partly compete with treatment for the same difference in slope, so flexible calendar adjustment may remove residual period confounding but may also absorb part of a treatment difference. Baseline EDSS was missing in 38.9% of participants, and single imputation may have limited both confounding control and the estimation of uncertainty.

Requiring a valid baseline SDMT, no earlier recorded testing, and at least two on-treatment assessments selected a subgroup that differed from excluded individuals (Supplementary Table S1) and may limit generalizability. These requirements also selected on continued treatment and testing. Shorter time to the last on-treatment SDMT with natalizumab than rituximab (median 41.5 vs. 55.2 months), together with potentially informative treatment discontinuation or cessation of testing, may have influenced the estimated trajectories, and the year-3 contrasts in Table 3 rest on a smaller share of the natalizumab arm.

Testing frequency was closely associated with treatment, so adjustment for cumulative test number cannot conclusively separate practice effects from biological treatment effects. Estimates also depend on the assumed (logarithmic) relationship between test exposure and performance. Finally, administration format and use of alternate forms were not recorded. Changes from written to oral testing and variable use of alternate forms may therefore have introduced measurement differences associated with clinic, calendar period, and treatment.

## Conclusions

The SDMT difference between natalizumab and rituximab was substantially attenuated after adjustment for cumulative test exposure, leaving a small residual difference of uncertain clinical importance. These findings highlight the need to consider testing frequency and practice effects when comparing cognitive outcomes between DMTs in real-world cohorts.

## Supporting information

All supplementary material including RECORD checklist

## Data Availability

Data are held within the Swedish MS Registry and are available to qualified researchers subject to registry governance and Swedish data-protection legislation.

## Acknowledgements

We thank the Swedish MS Registry and the participating neurology clinics across Sweden for prospectively collecting and maintaining the data used in this study. Claude (Anthropic) and GPT (OpenAI) were used to review the Stata code and to edit the manuscript text; TPC verified all changes.

## Declarations

### Conflicts of Interest

TPC and SAH report no competing interests. FP has received research grants from Denka, Janssen, Merck KGaA, Novartis, Pfizer and Sanofi, and fees for serving on data monitoring committees in clinical trials with Lundbeck and Roche, and is supported by the Swedish Research Council (grant no. 2023-02639), the Region Stockholm (grant no. FoUI-987565), the Swedish Brain Fund (grant no. FO2023-0336) and Erling Persson’s Foundation (grant no. 2024-0127). KAM receives research funding from the Swedish Research Council and ALF Medicine and has received speaker honoraria from Biogen (2022) and Sanofi-Aventis (2023). KF has served on scientific advisory boards for Alexion, Biogen, Janssen, Merck, Novartis, Takeda and Roche, and has received financial research support from Merck and Amicus.

### Funding

This study was supported by a grant from the Swedish Research Council (2023-02567) to K.F. The funder had no role in study design, data collection, analysis, interpretation, or the decision to submit.

### Author Contributions

TPC conceived and designed the study, conducted the analyses, and drafted the manuscript. SAH contributed to early data exploration as part of an MSc thesis project. FP and KF critically revised the manuscript for important intellectual content. KAM conceived the study, provided supervision, and critically revised the manuscript for important intellectual content. All authors approved the final version.

