## Supplementary material for "More tests, higher scores: Testing frequency and SDMT differences between natalizumab and rituximab": All supplementary material including RECORD checklist

**Affiliations:**

#### Supplementary Methods

*Baseline SDMT window and score distribution*

The baseline SDMT was drawn from a 0–90 day window before initiation, and most baseline tests fell well inside it: 2,395 of 2,458 individuals (97.4%) had an SDMT within 30 days of initiation and 2,434 (99.0%) within 60 days. The 18,300 on-treatment assessments spanned almost the whole possible range of the test (observed range 6 to 110 points; mean 60.1, SD 15.9). Scores above 90 were recorded for 862 assessments (4.7%), 841 of them among natalizumab-treated PwMS. Free-text comments at these visits frequently noted the tested individuals were completing the SDMT evaluation before the 90 seconds were up, so these scores reflect the length of the test rather than the limits of cognitive ability.

*Imputation of baseline EDSS and disease duration*

Baseline EDSS was missing for 956 of 2,458 individuals (38.9%) and disease duration for 52 (2.1%). Missingness differed by arm for both variables: baseline EDSS was missing for 40.6% of rituximab and 36.7% of natalizumab initiators ($p = 0.048$), and disease duration for 2.8% and 1.2% respectively ($p = 0.006$). Both were filled by a single stochastic draw conditional on age, sex, and region, with disease duration additionally conditioning the EDSS imputation. Neither imputation model contains treatment or SDMT, so imputed values carry no arm-specific or cognition-specific signal, and the imputed portion of baseline EDSS cannot contribute to confounding control.

*Time-specification comparison*

Linear and restricted cubic spline specifications were fitted unadjusted by maximum likelihood and compared by AIC. The splines used Harrell's recommended percentile placements: 3 knots at 10, 50, and 90; 4 knots at 5, 35, 65, and 95; and 5 knots at 5, 27.5, 50, 72.5, and 95. Because the candidate models share the same outcome, analytic observations, likelihood, and random-effects structure, AIC differences reflect the time specification alone. Only the AIC ranking is interpreted from that comparison; its standard errors are model-based rather than robust, and the reported trajectory models use robust standard errors throughout.

*Machine-learning separability diagnostic*

We fitted regularised gradient-boosted classifiers[1] of treatment across four feature sets: testing-process features (on-treatment SDMT count, testing rate, inter-test-interval summaries, and years to the first and last on-treatment test), baseline clinical indication (age, sex, EDSS, disease duration, treatment-naïve status, baseline SDMT), calendar year of initiation, and all features combined. Calendar year was kept separate from clinical indication to avoid inflating the indication signal. Five-fold cross-validated AUC summarised separability (Supplementary Table S4), and mean absolute SHAP values[2] from the combined model ranked feature contributions (Supplementary Table S5). Gradient-boosted trees were regularised (300 trees, maximum depth 3, learning rate 0.05, subsample fraction 0.8, minimum 20 observations per leaf, seed 20260602) to ensure out-of-sample generalisation and keep SHAP attributions interpretable. The diagnostic characterises the observed joint distribution of treatment and testing; it is not a propensity adjustment and does not correct the comparison. The classifiers used scikit-learn's gradient-boosting implementation (Python 3.14.0, scikit-learn 1.9.1).

**Supplementary Table S1.** Baseline characteristics by SDMT data availability among the 9,060 otherwise eligible individuals (2,458 included vs. 6,602 excluded).

| No. (Column %) or Median (Q1, Q3) | Total N=9,060 | SDMT-based inclusion N=2,458 | SDMT-based exclusion N=6,602 |
|---|---|---|---|
| Age at treatment initiation (years) | 37.0 (29.0, 46.0) | 35.0 (28.0, 44.0) | 37.0 (30.0, 46.0) |
| Sex | | | |
| Female | 6,392 (71) | 1,705 (69) | 4,687 (71) |
| Disease duration at treatment (years) | 3.0 (0.5, 9.2) | 2.4 (0.4, 7.8) | 3.2 (0.5, 9.7) |
| Disease duration | | | |
| <2 years | 3,911 (43) | 1,146 (47) | 2,765 (42) |
| 2-5 years | 1,555 (17) | 449 (18) | 1,106 (17) |
| >5 years | 3,594 (40) | 863 (35) | 2,731 (41) |
| Baseline EDSS | 2.0 (1.0, 2.6) | 2.0 (1.0, 2.6) | 1.9 (1.0, 2.5) |
| Treatment naive (no prior DMT) | 4,649 (51) | 1,391 (57) | 3,258 (49) |
| Duration of prior DMT exposure (months) | 0.0 (0.0, 42.4) | 0.0 (0.0, 25.8) | 1.0 (0.0, 49.3) |

*Footnote:* Sex was not recorded for fewer than 5 individuals, so only the female row is shown. EDSS, Expanded Disability Status Scale; SDMT, Symbol Digit Modalities Test; DMT, disease-modifying therapy; Q1 and Q3, first and third quartiles. Baseline EDSS and disease duration include values filled by single stochastic imputation (Supplementary Methods), so EDSS quartiles need not fall on EDSS steps.

**Supplementary Table S2.** Time-specification fit comparison (linear, RCS-3, RCS-4, RCS-5) by AIC.

| | Linear | | | RCS-3 | | | RCS-4 | | | RCS-5 | | |
|---|---|---|---|---|---|---|---|---|---|---|---|---|
| | **Coef.** | **95% CI** | **p-value** | **Coef.** | **95% CI** | **p-value** | **Coef.** | **95% CI** | **p-value** | **Coef.** | **95% CI** | **p-value** |
| Natalizumab (vs. Rituximab) at Baseline | 3.95 | (2.94, 4.95) | <0.001 | 0.67 | (-0.38, 1.73) | 0.21 | -0.01 | (-1.16, 1.15) | >0.99 | -0.73 | (-2.03, 0.57) | 0.27 |
| Years since Treatment Initiation | 0.31 | (0.18, 0.43) | <0.001 | 0.66 | (0.43, 0.89) | <0.001 | 1.91 | (1.43, 2.39) | <0.001 | 1.96 | (1.10, 2.82) | <0.001 |
| NTZ x Years (linear) | 0.50 | (0.31, 0.69) | <0.001 | 2.97 | (2.68, 3.27) | <0.001 | 4.07 | (3.50, 4.65) | <0.001 | 5.28 | (4.28, 6.27) | <0.001 |
| Time spline term (RCS-3) | | | | -0.68 | (-1.05, -0.31) | <0.001 | | | | | | |
| NTZ x Time spline term (RCS-3) | | | | -4.78 | (-5.22, -4.33) | <0.001 | | | | | | |
| Time spline term 2 (RCS-4) | | | | | | | -13.01 | (-16.95, -9.07) | <0.001 | | | |
| Time spline term 3 (RCS-4) | | | | | | | 22.70 | (15.64, 29.75) | <0.001 | | | |
| NTZ x Time spline term 2 (RCS-4) | | | | | | | -21.18 | (-25.93, -16.43) | <0.001 | | | |
| NTZ x Time spline term 3 (RCS-4) | | | | | | | 31.32 | (22.83, 39.81) | <0.001 | | | |
| Time spline term 2 (RCS-5) | | | | | | | | | | -16.82 | (-34.10, 0.47) | 0.057 |
| Time spline term 3 (RCS-5) | | | | | | | | | | 22.08 | (-11.07, 55.23) | 0.19 |
| Time spline term 4 (RCS-5) | | | | | | | | | | 1.07 | (-19.61, 21.75) | 0.92 |
| NTZ x Time spline term 2 (RCS-5) | | | | | | | | | | -58.61 | (-79.18, -38.04) | <0.001 |
| NTZ x Time spline term 3 (RCS-5) | | | | | | | | | | 95.56 | (55.83, 135.30) | <0.001 |
| NTZ x Time spline term 4 (RCS-5) | | | | | | | | | | -40.90 | (-66.07, -15.73) | 0.001 |
| Intercept | 53.43 | (52.75, 54.11) | <0.001 | 52.90 | (52.17, 53.63) | <0.001 | 51.67 | (50.83, 52.52) | <0.001 | 51.64 | (50.64, 52.64) | <0.001 |
| AIC | | 124099.97 | | | 122342.92 | | | 122066.72 | | | 122026.04 | |

*Footnote:* Fitted unadjusted by maximum likelihood on a common sample. Only the AIC ranking is interpreted; the coefficients and intervals shown carry model-based rather than robust standard errors, so they differ from the corresponding RCS-5 entries in Supplementary Table S3, which uses robust standard errors throughout. NTZ, natalizumab.

**Supplementary Table S3.** Full coefficient tables for the three sequentially adjusted five-knot spline models, including the common time slope, the spline and treatment-by-spline terms, covariates, variance components, and AIC.

| | Unadjusted | | | Adjusted | | | + Practice | | |
|---|---|---|---|---|---|---|---|---|---|
| | **Coef.** | **95% CI** | **p-value** | **Coef.** | **95% CI** | **p-value** | **Coef.** | **95% CI** | **p-value** |
| Natalizumab (vs. Rituximab) at Baseline | -0.73 | (-2.16, 0.70) | 0.32 | -3.30 | (-4.65, -1.94) | <0.001 | -1.69 | (-2.88, -0.50) | 0.005 |
| Years since Treatment Initiation | 1.96 | (1.17, 2.75) | <0.001 | 1.23 | (0.41, 2.06) | 0.003 | -2.72 | (-3.83, -1.62) | <0.001 |
| Time spline term 2 | -16.82 | (-33.19, -0.45) | 0.044 | -14.63 | (-30.86, 1.60) | 0.077 | 9.21 | (-7.64, 26.07) | 0.28 |
| Time spline term 3 | 22.08 | (-9.66, 53.82) | 0.17 | 19.00 | (-12.51, 50.51) | 0.24 | -16.81 | (-49.06, 15.45) | 0.31 |
| Time spline term 4 | 1.07 | (-19.40, 21.55) | 0.92 | 1.57 | (-18.82, 21.96) | 0.88 | 12.85 | (-7.65, 33.36) | 0.22 |
| NTZ x Years (linear) | 5.28 | (3.85, 6.71) | <0.001 | 5.05 | (3.66, 6.45) | <0.001 | 1.38 | (0.18, 2.57) | 0.024 |
| NTZ x Time spline term 2 | -58.61 | (-86.13, -31.10) | <0.001 | -58.78 | (-85.97, -31.59) | <0.001 | 0.08 | (-25.71, 25.88) | >0.99 |
| NTZ x Time spline term 3 | 95.56 | (42.84, 148.29) | <0.001 | 95.36 | (43.14, 147.58) | <0.001 | -8.83 | (-59.21, 41.55) | 0.73 |
| NTZ x Time spline term 4 | -40.90 | (-74.63, -7.18) | 0.017 | -40.25 | (-73.81, -6.70) | 0.019 | 11.84 | (-21.17, 44.85) | 0.48 |
| Age at treatment initiation (years) | | | | -0.14 | (-0.17, -0.11) | <0.001 | -0.15 | (-0.18, -0.11) | <0.001 |
| Female (vs. Male) | | | | 0.08 | (-0.57, 0.74) | 0.80 | 0.19 | (-0.46, 0.84) | 0.56 |
| Baseline EDSS | | | | -0.54 | (-0.78, -0.30) | <0.001 | -0.55 | (-0.78, -0.31) | <0.001 |
| Disease duration at treatment (years) | | | | -0.01 | (-0.05, 0.04) | 0.83 | 0.01 | (-0.03, 0.06) | 0.59 |
| Treatment naive (no prior DMT) | | | | 0.19 | (-0.55, 0.93) | 0.61 | 0.15 | (-0.58, 0.89) | 0.69 |
| Baseline SDMT score | | | | 0.81 | (0.77, 0.85) | <0.001 | 0.81 | (0.77, 0.85) | <0.001 |
| Calendar year spline term 1 (linear, centred) | | | | -0.49 | (-0.77, -0.21) | <0.001 | -0.17 | (-0.48, 0.13) | 0.27 |
| Calendar year spline term 2 | | | | 0.13 | (-0.16, 0.43) | 0.38 | -0.08 | (-0.39, 0.22) | 0.61 |
| Calendar year spline term 1 (linear, centred) # Years since Treatment Initiation | | | | -0.23 | (-0.32, -0.14) | <0.001 | -0.16 | (-0.24, -0.08) | <0.001 |
| Calendar year spline term 2 # Years since Treatment Initiation | | | | 0.26 | (0.15, 0.38) | <0.001 | 0.24 | (0.13, 0.35) | <0.001 |
| Health care region (sparse regions pooled) | | | | | | | | | |
| Norra | | | | | *Reference* | | | *Reference* | |
| Stockholms region | | | | 0.00 | (-0.90, 0.90) | >0.99 | 0.58 | (-0.31, 1.46) | 0.20 |
| Sydöstra | | | | -0.39 | (-1.77, 0.99) | 0.58 | 0.24 | (-1.09, 1.57) | 0.73 |
| Södra | | | | 1.54 | (0.44, 2.65) | 0.006 | 1.55 | (0.50, 2.59) | 0.004 |
| Uppsala-Örebro | | | | 0.02 | (-1.12, 1.17) | 0.97 | 0.46 | (-0.67, 1.58) | 0.43 |
| Västsvenska | | | | 3.81 | (2.71, 4.91) | <0.001 | 3.14 | (2.01, 4.27) | <0.001 |
| Ln(test number) | | | | | | | 6.60 | (5.35, 7.86) | <0.001 |
| Intercept | 51.64 | (50.75, 52.53) | <0.001 | 15.78 | (12.82, 18.74) | <0.001 | 14.41 | (11.45, 17.37) | <0.001 |
| Variance: Patient identifier (Years since Treatment Initiation) | 2.05 | (1.58, 2.65) | | 1.92 | (1.49, 2.47) | | 1.76 | (1.37, 2.27) | |
| Variance: Patient identifier (Intercept) | 138.54 | (128.56, 149.29) | | 44.73 | (39.68, 50.43) | | 46.72 | (41.00, 53.23) | |
| Covariance: Patient identifier (Years since Treatment Initiation, Intercept) | -0.21 | (-1.63, 1.22) | | -0.82 | (-1.76, 0.11) | | -1.61 | (-2.63, -0.59) | |
| Residual Variance | 26.36 | (23.97, 29.00) | | 26.36 | (23.98, 28.98) | | 25.26 | (23.10, 27.63) | |
| AIC | | 122026.04 | | | 119505.46 | | | 118743.75 | |

*Footnote:* The test-number coefficient is the change in SDMT points per one-unit increase in the natural log of cumulative test number; multiply by 0.693 (ln 2) for the change per doubling of test number. NTZ, natalizumab.

**Supplementary Table S4.** Cross-validated treatment-classification performance by feature set (testing process, baseline clinical indication, calendar year, all combined).

| Model | Method | Task | Evaluated on | N | Accuracy | F1 | AUC |
|---|---|---|---|---|---|---|---|
| Testing process | Boost | Classification | Cross-validation | 2,458 | 0.8206 | 0.7843 | 0.8839 |
| Clinical indication | Boost | Classification | Cross-validation | 2,458 | 0.6310 | 0.5522 | 0.6775 |
| Calendar year | Boost | Classification | Cross-validation | 2,458 | 0.7787 | 0.7194 | 0.8244 |
| All features | Boost | Classification | Cross-validation | 2,458 | 0.8588 | 0.8328 | 0.9224 |

*Footnote:* Regularised gradient boosting, five-fold cross-validation; accuracy, F1, and AUC. AUC near 1 indicates that the feature set nearly determines treatment; AUC near 0.5 indicates chance-level discrimination and does not by itself establish exchangeability.

**Supplementary Table S5.** Mean absolute SHAP values from the combined classifier, ranking testing-process, clinical, and calendar features as predictors of treatment assignment.

| Rank | Feature | Mean \|SHAP\| |
|---|---|---|
| 1 | Calendar year of initiation (centered) | 1.1209 |
| 2 | Years to first on-treatment SDMT | 0.5287 |
| 3 | Min inter-test gap (yr) | 0.5066 |
| 4 | Age at treatment initiation (years) | 0.3853 |
| 5 | Mean inter-test gap (yr) | 0.3405 |
| 6 | Years to last SDMT | 0.3244 |
| 7 | SDMT testing rate (tests/yr) | 0.3238 |
| 8 | Disease duration at treatment (years) | 0.2233 |
| 9 | On-treatment SDMT count | 0.1151 |
| 10 | Treatment naive (no prior DMT) | 0.0905 |
| 11 | Baseline SDMT score | 0.0826 |
| 12 | Baseline EDSS | 0.0747 |
| 13 | SD of inter-test gap (yr) | 0.0568 |
| 14 | Female (vs. Male) | 0.0086 |

**Supplementary Table S6.** Predicted SDMT with 95% confidence intervals at cumulative test numbers 2 to 10 by treatment group, the values plotted in Supplementary Figure S3B.

| SDMT test number | Rituximab, predicted SDMT | Rituximab, 95% CI | Natalizumab, predicted SDMT | Natalizumab, 95% CI |
|---|---|---|---|---|
| 2 | 52.59 | (51.52, 53.67) | 51.71 | (50.32, 53.10) |
| 3 | 54.39 | (53.63, 55.15) | 54.73 | (53.80, 55.65) |
| 4 | 55.66 | (55.02, 56.30) | 56.87 | (56.21, 57.52) |
| 5 | 56.65 | (56.00, 57.29) | 58.53 | (57.97, 59.09) |
| 6 | 57.46 | (56.74, 58.17) | 59.88 | (59.30, 60.47) |
| 7 | 58.14 | (57.33, 58.95) | 61.03 | (60.35, 61.71) |
| 8 | 58.73 | (57.82, 59.64) | 62.02 | (61.23, 62.82) |
| 9 | 59.25 | (58.25, 60.26) | 62.90 | (61.99, 63.82) |
| 10 | 59.72 | (58.62, 60.81) | 63.69 | (62.65, 64.72) |

*Footnote:* Predictions from the model with a treatment-by-log-test-number interaction, with time since initiation and baseline covariates at their means, region as observed, and robust standard errors.

**Supplementary Figure S1.** CONSORT-style flow diagram of cohort selection from the Swedish MS Registry, 2010–2024. NTZ, natalizumab; RTX, rituximab; RRMS, relapsing-remitting MS; SDMT, Symbol Digit Modalities Test; “No baseline”, no baseline SDMT.

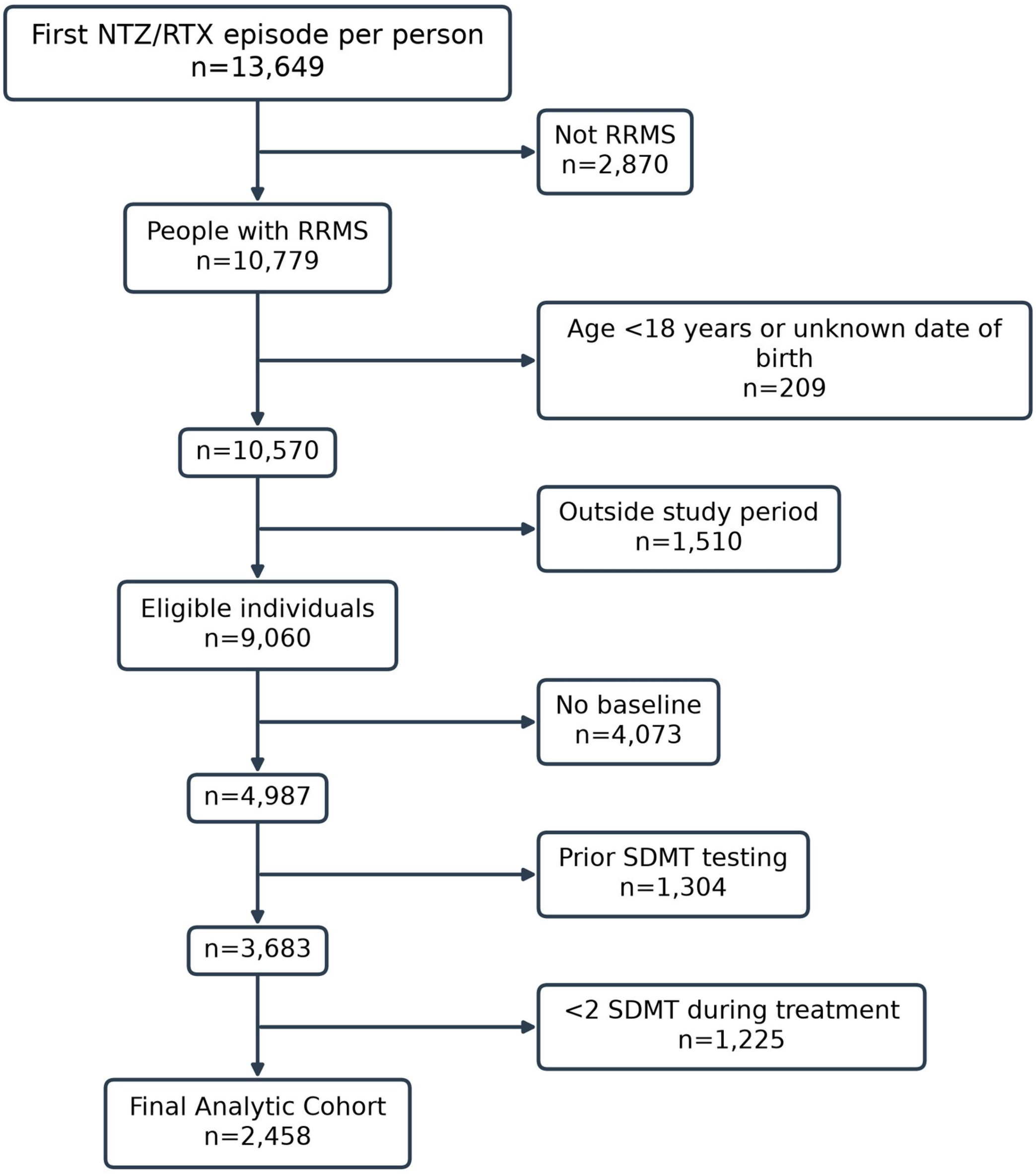

**Supplementary Figure S2.** Time specification diagnostic: lowess of raw SDMT data overlaid with linear, RCS-3, RCS-4, and RCS-5 fitted trajectories over 0–5 years, in (A) rituximab and (B) natalizumab initiators.

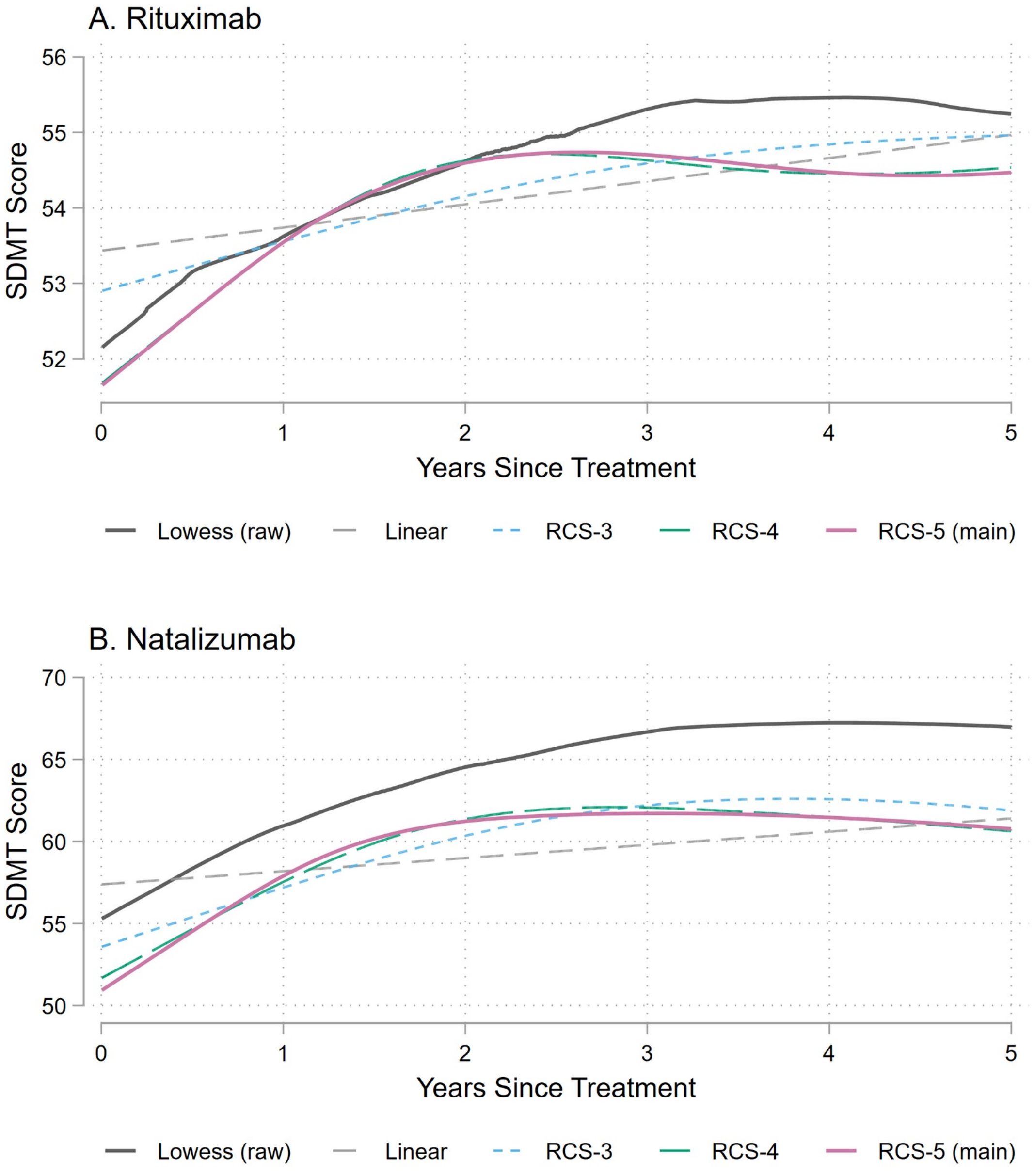


The relevant comparison is of curve shape, not level. The lowess weights every assessment equally, so it is pulled towards the most frequently tested patients, whereas the mixed models weight patients equally through the random intercept. In the natalizumab panel, where testing is densest, the lowess therefore sits several points above every fitted specification.

**Supplementary Figure S3.** Model-predicted longitudinal SDMT trajectories by treatment group.

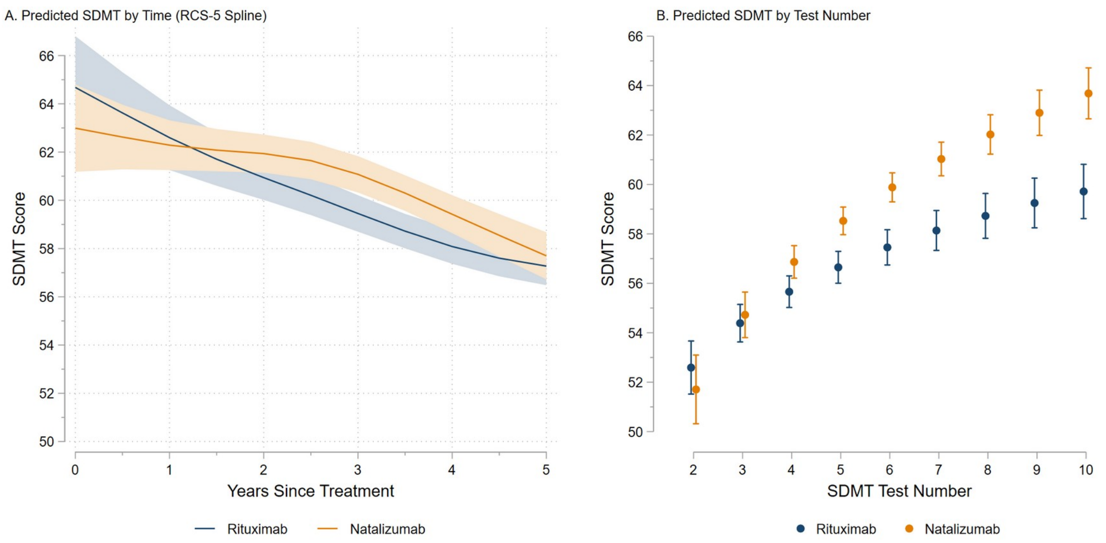


(A) Predicted SDMT over time from the primary five-knot spline model, holding cumulative test number at its sample mean across all time points; because testing is held fixed, both arms decline rather than improve, and the between-arm contrasts at 1, 2, and 3 years are those in Table 3. (B) Predicted SDMT at each cumulative test number from 2 to 10 from a separate model that replaces the treatment-by-time interaction with a treatment-by-log-test-number interaction, holding time since initiation at its sample mean; the plotted values are listed in Supplementary Table S6. Both panels adjusted for age, sex, baseline SDMT, baseline EDSS, disease duration, treatment-naïve status, calendar year of initiation (restricted cubic spline, interacted with time), and health care region. Because covariates are held at their means and region as observed, absolute levels at early follow-up are extrapolations, and only the between-arm contrast and trajectory shape are interpretable. Equal-test-number contrasts in panel B rely on the few individuals with comparable test counts in both arms and are not treatment-effect estimates. Shaded bands (A) and whiskers (B) indicate 95% confidence intervals.

### RECORD checklist

**Manuscript:** More tests, higher scores: Testing frequency and SDMT differences between natalizumab and rituximab

Benchimol EI, et al. PLoS Med 2015;12(10):e1001885 (STROBE cohort items with RECORD extensions). Items not reported or not applicable are noted. Methods subsections: Study design and setting, Study population, Exposure (Exposure, outcome, and follow-up), Practice (Practice effects and test-number adjustment), Covariates, Statistical analysis, Ethics. SM, Supplementary Methods; S, Supplementary Table; Fig S, Supplementary Figure.

| Item | Topic | Where reported |
|---|---|---|
| 1 | Design in title; informative abstract | Abstract (design not named in title or abstract) |
| 1.1 | Data type and databases named | Abstract, Methods; Methods, Study design and setting (not in title) |
| 1.2 | Region and time frame | Abstract, Methods (not in title) |
| 1.3 | Linkage stated | Not applicable: single registry, no linkage described |
| 2 | Background and rationale | Introduction |
| 3 | Objectives | Abstract, Objective; Introduction, last sentence |
| 4 | Study design | Methods, Study design and setting |
| 5 | Setting and dates | Methods, Study design and setting, Study population, Exposure |
| 6 | Eligibility, follow-up | Methods, Study population, Exposure; Fig S1 |
| 6.1 | Codes and algorithms for the population | Methods, Study population (registry fields described in words; no variable list or algorithm) |
| 6.2 | Validation of codes | Methods, Study design and setting (registry chart-review validation; no validation of the SDMT or DMT-episode fields) |
| 6.3 | Linkage flow diagram | Not applicable (no linkage); cohort flow in Fig S1 |
| 7 | Variable definitions | Methods, Exposure, Practice, Covariates, Statistical analysis; Table 3 footnote |
| 7.1 | Codes for all variables | Not reported (no registry variable names or code list) |
| 8 | Data sources and measurement | Methods, Study design and setting, Exposure |
| 9 | Bias | Methods, Practice; Discussion, Strengths and limitations |
| 10 | Study size | Results, Cohort and baseline characteristics; Fig S1 (no sample-size calculation) |

| Item | Topic | Where reported |
|---|---|---|
| 11 | Quantitative variables | Methods, Practice, Covariates, Statistical analysis; SM, Time-specification comparison |
| 12 | Statistical methods, interactions, missing data, follow-up, sensitivity analyses | Methods, Covariates, Statistical analysis; SM; S2, S3; alternative analyses: time specifications (S2, Fig S2), equal-test-number predictions (Fig S3, S6), observed testing schedules (Figure 2) (no method for loss to follow-up or informative discontinuation; no sensitivity analyses of exposure, outcome or imputation definitions) |
| 12.1 | Data access | Methods, Ethics; Declarations, Data Availability (extent of investigators' access to the registry not stated) |
| 12.2 | Data cleaning | Methods, Exposure, Statistical analysis; SM, Baseline SDMT window and score distribution, Imputation of baseline EDSS and disease duration |
| 12.3 | Linkage method and quality | Not applicable (no linkage) |
| 13, 13.1 | Participants at each stage; selection | Results, Cohort and baseline characteristics; Fig S1; S1 |
| 14 | Descriptive data, missing data, follow-up time | Table 1; Results, Cohort and baseline characteristics; SM, Imputation of baseline EDSS and disease duration |
| 15 | Outcome data | Table 2; Results, Unadjusted SDMT outcomes |
| 16 | Main results, category boundaries, absolute measures | Results, Longitudinal trajectories depend on the practice-effect specification; Table 3; Figure 2; S3 |
| 17 | Other analyses | Results, Longitudinal trajectories, Prediction of treatment assignment; Figure 1; S2, S4 to S6; Fig S2, S3 |
| 18 | Key results | Discussion, first paragraph |
| 19, 19.1 | Limitations; implications of routinely collected data | Discussion, Strengths and limitations |
| 20 | Interpretation | Discussion; Conclusions |
| 21 | Generalisability | Discussion, Strengths and limitations |
| 22 | Funding | Declarations, Funding |
| 22.1 | Protocol, data and code access | Declarations, Data Availability (no protocol registration or code access stated) |
